# Clinical epidemiology of twin deliveries in The Gambia and Burkina Faso: Secondary analyses of PregnAnZI-2 clinical trial data

**DOI:** 10.1101/2025.10.25.25338790

**Authors:** Joquina Chiquita Jones, Toussaint Rouamba, Joel D. Bognini, Bully Camara, Usman N Nakakana, Athanase M. Somé, Nathalie Beloum, Guétawendé J W Nassa, Fatoumata Sillah, Edmond Yabré Sawadogo, Madikoi Danso, Shashu Graves, Bai Lamin Dondeh, Diagniagou Lankoandé, Ebrahim Ndure, Yusupha Njie, Christian Bottomley, Umberto d’Alessandro, Halidou Tinto, Anna Roca, Helen Brotherton

## Abstract

**Introduction:** Twins have higher risk of adverse clinical outcomes in all settings, but especially in low- and middle-income countries. This study aims to understand the clinical epidemiology of twins from varied West African settings, focusing on prevalence and determinants of twins, and adverse perinatal outcomes associated with twin deliveries. These context specific insights may help to inform public health and clinical approaches to improving health and survival of this vulnerable newborn group.

**Methods:** We conducted secondary analyses from the PregnAnZI-2 randomised clinical trial, which included data from pregnant women and their offspring who were delivered at eight peripheral health facilities in rural Burkina Faso and two urban health facilities in The Gambia. Logistic regression was used to identify associations between primary outcome (twin deliveries) and both risk factors and adverse outcomes and was informed by a novel conceptual framework.

**Results:** Data from 12,192 newborns born from 11,983 women were analysed. The average twinning rate was 35 per 1000 total births (95% CI: 32-39): 27 per 1000 total births (95% CI: 23-31) in The Gambia and 45 per 1000 total births (95% CI: 40-51) in Burkina Faso. Risk factors for twinning included maternal ethnicity and parity. Twins had increased risk of low birth weight (OR =17.2, 95% CI:13.90-21.30, P= <0.001), low 5-min Apgar score (proxy for intrapartum related asphyxia) (OR =4.28, 95% CI: 2.79-6.33, P <0.001), intrapartum stillbirth (OR=4.69, 95% CI:2.47-8.22, P<0.001), and hospitalisation during the neonatal period (OR =1.69 95% CI:1.10-2.47, P= 0.011). Neonatal mortality was four-fold higher for twins than singletons (OR =4.23, 95% CI: 2.40-7.50, P <0.001), and among twin deaths, the second twin was more likely to die compared to the first twin (OR=2.46, 95% CI :1.05-7.59, P=0.048).

**Conclusion:** Twins are an important contributor to stillbirth and neonatal morbidity and mortality in West Africa. Efforts to identify twin pregnancies early, with optimisation of antenatal and intrapartum management could improve outcomes for this vulnerable newborn group.

**Trial Registration:** NCT03199547: Clinicaltrials.gov. Registered on 23rd June 2017

## INTRODUCTION

Worldwide, twin births occur in 1 in 76 pregnancies, equivalent to 13.1 per 1000 live births.(1) Sub-Saharan Africa (SSA) has the highest rates of twin births globally, (1, 2) with significant regional variation (3) ranging from >18 per 1000 live births in West and Central Africa to 11-15 per 1000 live births in Southern African countries.(1) Twin pregnancies are associated with a substantially higher risk of antenatal and perinatal adverse outcomes, including preterm birth, low birth weight (LBW), stillbirth, intrapartum-related asphyxia (IRA), and neonatal mortality, compared to singleton pregnancies.(4, 5) As a consequence, surviving twins face an increased risk of long-term morbidities and neurodevelopmental impairment(6), with huge impact on individuals, families, and societies.

Generic risk factors for twin pregnancies are well established and include maternal characteristics such as older age, multiparity, family history of twinning, and nutritional status, as well as lifestyle factors such as smoking.(5, 7, 8, 9) Other influences include socioeconomic status, assisted reproductive therapy, and race/ethnicity.(1, 10, 11) However, specific determinants for twinning within West African populations remain poorly explored, despite their importance in guiding targeted public health strategies aimed at early identification of twin pregnancies and optimising maternity care. This is particularly important if West Africa, a sub-region with both high twinning rates and the highest global neonatal mortality rates, is to achieve the sustainable development goal target of reducing avoidable neonatal mortality by 2030.(12) Understanding the clinical epidemiology of this group of vulnerable newborns in West Africa is crucial to delivering appropriate antenatal and intrapartum care to women with twin pregnancies in-order to improve twin outcomes.(5, 13) This study aims to examine the clinical epidemiology of twins from varied settings in West Africa. Specifically, we aim to determine the prevalence of twin births, identify risk factors associated with twinning, and quantify the adverse perinatal outcomes associated with twins.

## METHODS

### Study design

This is a secondary data analysis of the PregnAnZI-2 trial (clinicaltrials.gov ref: NCT03199547), which was a phase III, double-blind, placebo-controlled randomised clinical trial conducted in The Gambia and Burkina Faso between 2017 and 2021. Intra-partum azithromycin or placebo were administered to women in labour to assess the effect on a composite primary outcome of neonatal mortality or infection.(14) The whole trial cohort was included in this secondary study as the intervention was not associated with reductions in neonatal mortality or sepsis and would not influence twinning rates due to intrapartum timing of administration.(15)

### Setting

The PregnAnZI-2 trial was conducted at two urban government health facilities in The Gambia: Bundung Maternal Child Health Hospital (BMCHH) and Serekunda Health Centre (SHC). The two sites manage approximately 7000 deliveries per year.(14) Approximately 84% of deliveries in The Gambia occur at health facilities.(16) At the time of data collection, SHC lacked the capacity to provide emergency obstetric care and women requiring an emergency caesarean section were referred to the Kanifing General Hospital, ∼3.4 kilometres (Km) distance away. BMCHH provided emergency obstetric care and had a neonatal unit capable of providing WHO level 2 small and sick newborn care. Women experiencing complications of labour and unwell neonates were occasionally referred to the tertiary level obstetric department and national neonatal referral unit at the Edward Francis Small Teaching Hospital in Banjul, ∼15.5 Km distance away.

In Burkina Faso, women were recruited at eight peripheral health facilities in the districts of Nanoro and Yako. Approximately 94% of deliveries in Burkina Faso occur at health facilities.(17) The eight combined health facilities conducted ∼3800 deliveries annually, with antenatal, intrapartum and postnatal care available at primary care level. All mothers and newborns requiring specialised obstetric or neonatal hospital care were referred to the Saint Camille hospital in CRUN (25 Km average distance). The neonatal mortality rate (NMR) at study onset was the same in both countries (25/1000 live births), (18) with stillbirth rates of 21 per 1000 total births in The Gambia and 20 per 1000 total births in Burkina Faso.(19)

### Participants

Participants were pregnant women ≥ 16 years old who consented to take part in the PregnAnZI-2 trial and attended a study site for delivery. Women were excluded if they had a known acute or chronic condition (including HIV infection), planned caesarean section, severe congenital malformation detected in-utero, intra-uterine death, macrolide allergy or had taken a drug known to prolong QT interval during the preceding 2 weeks.(20) All offspring (stillbirths and livebirths) of recruited women were included in this secondary analysis. Assisted conception methods (e.g., *In-vitro* fertilisation) were not available at the study sites or governmental health facilities in either country at the time of study, so participants are assumed to all have spontaneous conceptions.

### Outcome measures

The primary outcome was a binary variable (yes/no) for whether a newborn was a twin. Twins were compared to singletons in our analyses, with the exclusion of triplet deliveries (three newborns). Due to the absence of routine antenatal scanning in our setting, it was not possible to determine twin or triplet pregnancies in which an in-utero demise occurred prior to enrolment, hence twin deliveries instead of twin pregnancies were considered.

### Study procedures

Pregnant women provided written informed consent during antenatal care, which was verbally confirmed during labour and prior to enrolment. Trained research nurses collected socio-demographic and clinical data prospectively as soon as possible after enrolment using the antenatal card, partograph and delivery notes as source documents.(20) Data of interest included: maternal obstetric history; intrapartum events and delivery details; and perinatal outcome of stillbirth or live birth. Stillbirths were all classed as intrapartum in timing as women were only enrolled following confirmation of labour and measurement of foetal heart rate using a hand-held foetal doppler. Newborns who were unable to sustain breathing at delivery were resuscitated by health facility staff according to the Helping Babies Breathe protocol, with Apgar scores prospectively determined by health facility staff. Stillbirths were assigned by health facilities staff using clinical criteria (lack of breathing and heart rate) and verified by research staff. Birthweight was measured using calibrated digital scales, and easily recognisable congenital malformations were determined by trained research clinicians prior to discharge with post-hoc classification as per ICD-11 system.(21) Birth outcomes for each twin partner were independently recorded.

Active and passive surveillance were conducted to collect data on health outcome. All mothers and newborns underwent a detailed clinical assessment by a trained research clinician or paediatrician 4-24h after delivery and prior to health facility discharge. Discharge or referral to other health facilities was at the discretion of the health facility staff. Between discharge and 28 postnatal days, mothers were encouraged to contact the trial clinical team or visit a study health facility if they or their babies were unwell (passive surveillance), with travel expenses to encourage attendance for in-person review by the research team. In The Gambia, a trained research nurse conducted a home visit at day 28 to collect morbidity and mortality data and give clinical support when necessary (active surveillance). In Burkina Faso, women and their infants attended study health facilities at 28 days, with home visits by a field worker in the event of non-attendance. The decision to admit mothers and newborns during the follow-up period was made by trained research clinicians or paediatricians, based on their clinical judgement.

### Data management and statistical analysis

Data were collected electronically using encrypted mobile and computer devices at all study sites and uploaded to a study specific Research Electronic Data capture (REDCap) database. Data consistency checks and validation were conducted as per PregnANZI-2 trial procedures.(14) All analyses were conducted with Stata version 17.0 (Statacorp). Descriptive analyses were performed to determine the prevalence rates for all independent variables and adverse outcomes for the total cohort, with stratification according to plurality (singleton versus twin delivery). Twinning rates were computed by dividing the number of twin births by the total number of births and multiplying the outcome by 1000. We developed a novel conceptual framework based on biotoxicological plausibility and existing evidence for risk factors for twin pregnancy and impact of twinning on delivery outcome (Figure 1). This framework was used to select independent variables for logistic regression to identify pre-conception and conception risk factors for twins versus singletons, expressed as crude odds ratios, with 95% confidence intervals to represent degree of certainty, and *p* values. For categorical variables, the baseline category was chosen to reflect either a “normal pregnancy and delivery” (e.g., normal birth weight, non-extremes of parity) or the most prevalent grouping (e.g., ethnicity). An adjusted logistic regression model was developed for each independent variable found to have p<0.2 on unadjusted regression. In-order to avoid over adjusting for multiple variables on the same causal pathway (22), each model included only variables occurring before or at the same timepoint as the independent variable of interest. As ethnicity and country are co-linear with no overlap, we chose to adjust for ethnicity only to provide more granular insights to risk factors for twinning. Statistical significance was defined as p<0.05.

**Figure 1.**
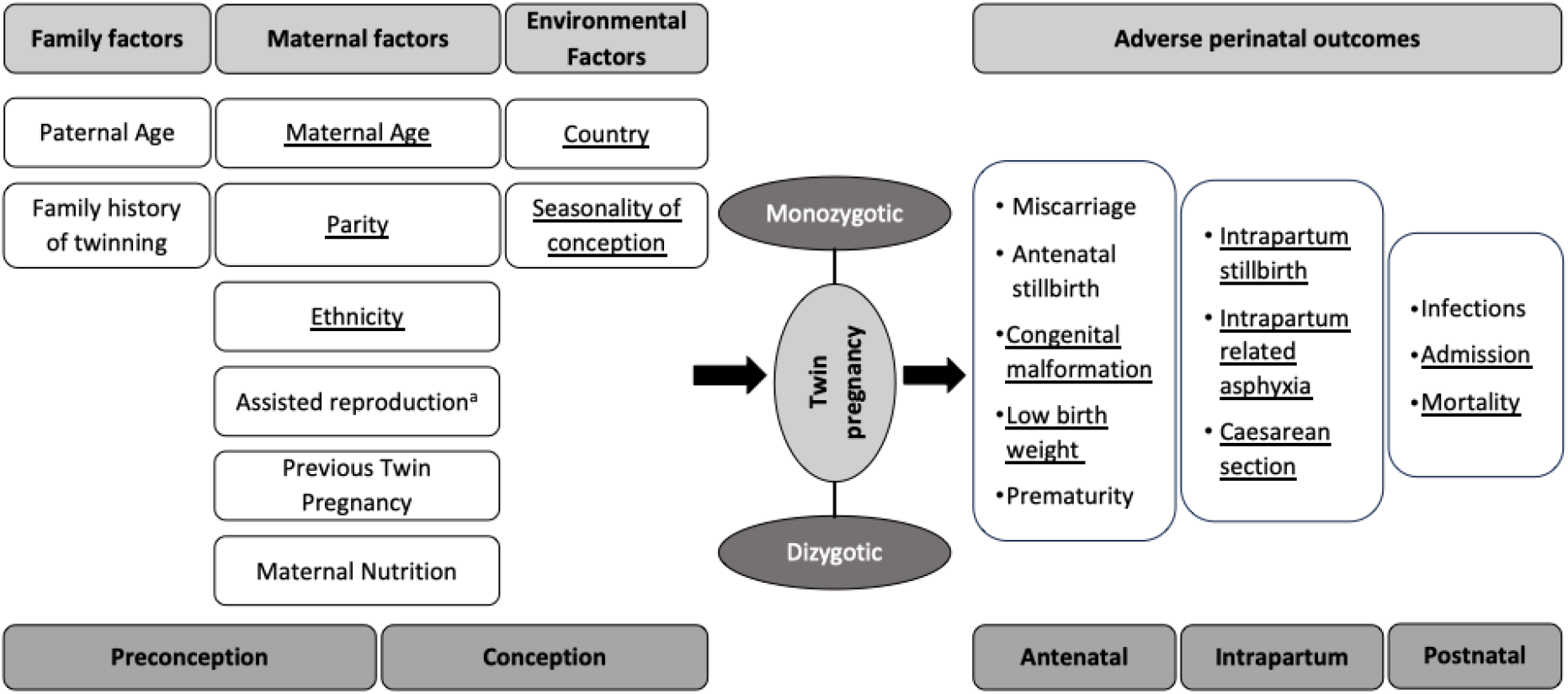
Conceptual Framework to understand development of twinning and impact of twin pregnancies on perinatal outcomes. Underlined variables indicate data available for analysis in this study Assisted reproduction methods including pre-conception therapies and invitro fertilization

## RESULTS

In total, 11,983 pregnant women and their 12,192 offspring were included in the analyses: 3.5% (428/12,192) were twins and 96.5% (11,764/12,192) were singletons (Figure 2). Overall, the average twinning rate for the two countries was 35 per 1000 total births (95% CI: 32-39): 27 per 1000 total births (95% CI: 23-31) in The Gambia and 45.4 per 1000 total births (95% CI: 40-51) in Burkina Faso (p<0.001) (Supplementary Table 1). Compared to women with a parity of 2, twins were more likely to be born to women with parity of 3 (aOR 1.50, 95% CI 1.08-2.11), or ≥4 (aOR 1.66, 95% CI 1.24-2.23), and primiparous women had reduced odds of twin birth (aOR 0.30, 95% CI 0.17-0.51). Younger maternal age (aOR 0.46, 95% CI 0.21-0.89) was associated with reduced odds of twins compared to women aged 20-35yrs. Maternal ethnicity of Gurunsi was associated with increased odds of having a twin compared to Mossi (aOR 1.69, 95% CI 1.09-2.62), with other ethnicities associated with reduced odds of twinning, such as Fula (aOR 0.55, 95% CI 0.35-0.81), Jola (aOR 0.54, 95% CI 0.33-0.84) and Mandinka (aOR 0.61, 95% CI 0.46-0.79) (Table 1). Overall, 14.3% (13/91) of intrapartum stillbirths and 12.7% (18/142) of neonatal deaths occurred in twins. Out of the twins who were either stillborn or died, 26% (8/31) were twin 1 and 74% (23/31) were twin 2 (cOR 3.1, 95% CI 1.41-7.55, *p*=0.007). There was a 4-fold higher odds of intrapartum stillbirth (cOR 4.69, 95% CI: 2.47-8.22, *p*<0.001) and neonatal mortality (cOR 4.23, 95% CI 2.55-5.70, *p*<0.001) for twins compared to singletons (Table 2). 21.9% (253/1156) of all LBW neonates in the cohort were twins and, compared to singletons, twins had 17-fold higher odds of being LBW (cOR 17.22, 95% CI 14.05-21.16, *p*<0.001). Twins were also more likely to have a low Apgar score at 1 minute (cOR 1.78, 95% CI 1.17-2.59, *p*=0.004) and 5 minutes (cOR 4.28, 95% CI: 2.79-6.33, *p*<0.001), be born by caesarean section (cOR 1.88, 95% CI 1.08-3.05, *p*= 0.016), and be admitted to hospital during the neonatal period (cOR 1.69, 95% CI 1.10-2.47, *p*=0.011).

**Table 1.**
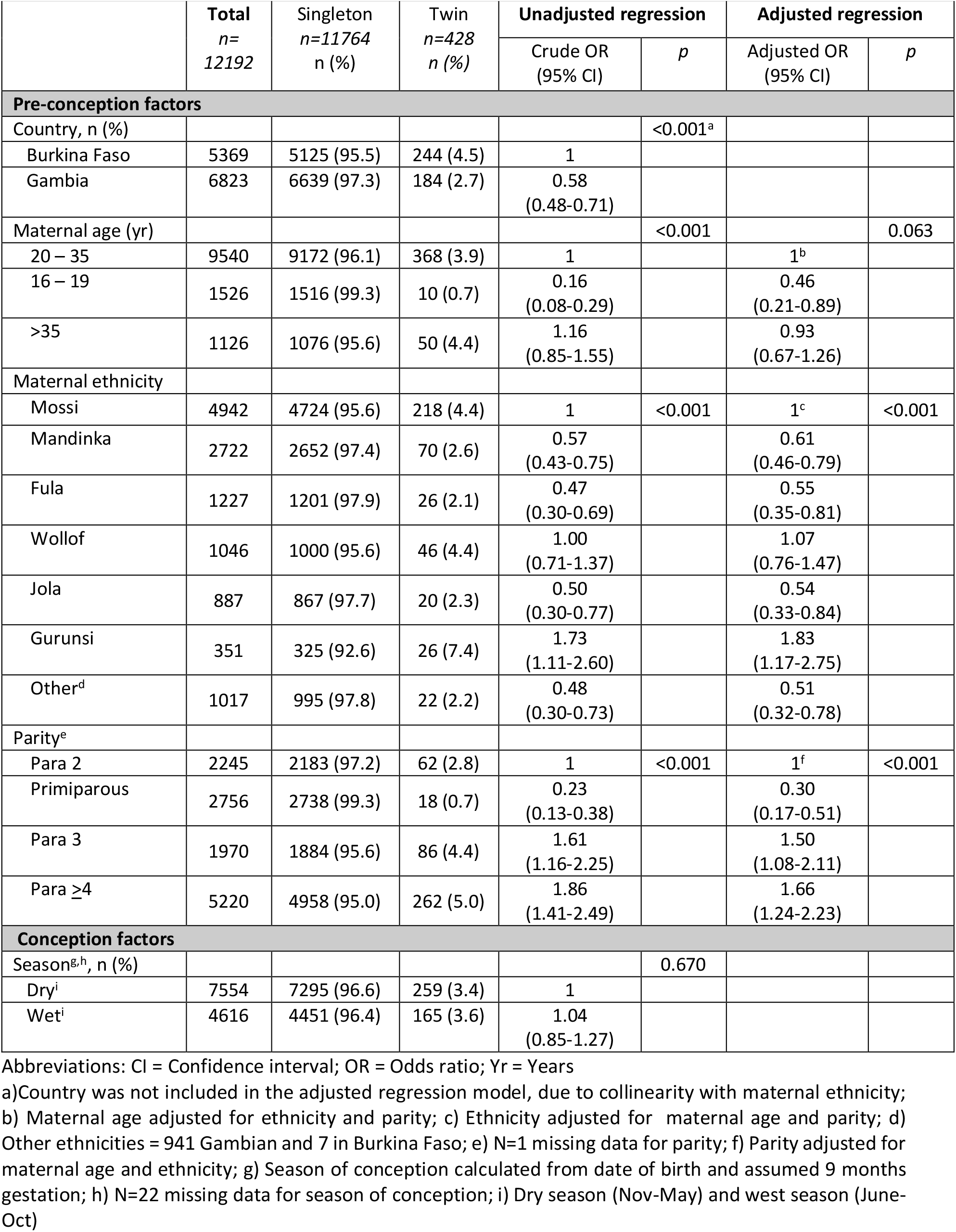
Pre-conception and conception factors associated with twinning in The Gambia and Burkina Faso.

|  | Total<br>n=<br>12192 | Singleton<br>n=11764<br>n (%) | Twin<br>n=428<br>n (%) | Unadjusted regression |  | Adjusted regression |  |
| --- | --- | --- | --- | --- | --- | --- | --- |
|  |  |  |  | Crude OR<br>(95% CI) | p | Adjusted OR<br>(95% CI) | p |
| Pre-conception factors |  |  |  |  |  |  |  |
| Country, n (%) |  |  |  |  | <0.001<sup>a</sup> |  |  |
| Burkina Faso | 5369 | 5125 (95.5) | 244 (4.5) | 1 |  |  |  |
| Gambia | 6823 | 6639 (97.3) | 184 (2.7) | 0.58<br>(0.48-0.71) |  |  |  |
| Maternal age (yr) |  |  |  |  | <0.001 |  | 0.063 |
| 20 – 35 | 9540 | 9172 (96.1) | 368 (3.9) | 1 |  | 1<sup>b</sup> |  |
| 16 – 19 | 1526 | 1516 (99.3) | 10 (0.7) | 0.16<br>(0.08-0.29) |  | 0.46<br>(0.21-0.89) |  |
| >35 | 1126 | 1076 (95.6) | 50 (4.4) | 1.16<br>(0.85-1.55) |  | 0.93<br>(0.67-1.26) |  |
| Maternal ethnicity |  |  |  |  |  |  |  |
| Mossi | 4942 | 4724 (95.6) | 218 (4.4) | 1 | <0.001 | 1<sup>c</sup> | <0.001 |
| Mandinka | 2722 | 2652 (97.4) | 70 (2.6) | 0.57<br>(0.43-0.75) |  | 0.61<br>(0.46-0.79) |  |
| Fula | 1227 | 1201 (97.9) | 26 (2.1) | 0.47<br>(0.30-0.69) |  | 0.55<br>(0.35-0.81) |  |
| Wollof | 1046 | 1000 (95.6) | 46 (4.4) | 1.00<br>(0.71-1.37) |  | 1.07<br>(0.76-1.47) |  |
| Jola | 887 | 867 (97.7) | 20 (2.3) | 0.50<br>(0.30-0.77) |  | 0.54<br>(0.33-0.84) |  |
| Gurunsi | 351 | 325 (92.6) | 26 (7.4) | 1.73<br>(1.11-2.60) |  | 1.83<br>(1.17-2.75) |  |
| Other<sup>d</sup> | 1017 | 995 (97.8) | 22 (2.2) | 0.48<br>(0.30-0.73) |  | 0.51<br>(0.32-0.78) |  |
| Parity<sup>e</sup> |  |  |  |  |  |  |  |
| Para 2 | 2245 | 2183 (97.2) | 62 (2.8) | 1 | <0.001 | 1<sup>f</sup> | <0.001 |
| Primiparous | 2756 | 2738 (99.3) | 18 (0.7) | 0.23<br>(0.13-0.38) |  | 0.30<br>(0.17-0.51) |  |
| Para 3 | 1970 | 1884 (95.6) | 86 (4.4) | 1.61<br>(1.16-2.25) |  | 1.50<br>(1.08-2.11) |  |
| Para ≥4 | 5220 | 4958 (95.0) | 262 (5.0) | 1.86<br>(1.41-2.49) |  | 1.66<br>(1.24-2.23) |  |
| Conception factors |  |  |  |  |  |  |  |
| Season<sup>g,h</sup>, n (%) |  |  |  |  | 0.670 |  |  |
| Dry<sup>i</sup> | 7554 | 7295 (96.6) | 259 (3.4) | 1 |  |  |  |
| Wet<sup>i</sup> | 4616 | 4451 (96.4) | 165 (3.6) | 1.04<br>(0.85-1.27) |  |  |  |

Abbreviations: CI = Confidence interval; OR = Odds ratio; Yr = Years
a) Country was not included in the adjusted regression model, due to collinearity with maternal ethnicity;
b) Maternal age adjusted for ethnicity and parity; c) Ethnicity adjusted for maternal age and parity; d)
Other ethnicities = 941 Gambian and 7 in Burkina Faso; e) N=1 missing data for parity; f) Parity adjusted for
maternal age and ethnicity; g) Season of conception calculated from date of birth and assumed 9 months
gestation; h) N=22 missing data for season of conception; i) Dry season (Nov-May) and wet season (June-
Oct)

**Table 2.** Adverse perinatal outcomes associated with twin deliveries at health facilities in The Gambia and Burkina Faso.

|  | Total<br>N=12,192 | Singleton<br>N=11,764 | Twin<br>N=428 | Crude Odds ratio<br>(95% CI) | *p* |
| --- | --- | --- | --- | --- | --- |
| **Intrapartum outcomes** |  |  |  |  |  |
| **Delivery mode<sup>a</sup>** |  |  |  |  | 0.016 |
| Vaginal | 11,936 | 11524 (96.5) | 412 (3.5) | 1 |  |
| Caesarean section | 254 | 238 (93.7) | 16 (6.3) | 1.88 (1.08-3.05) |  |
| **Low 1 min Apgar score (<7)<sup>b</sup>** |  |  |  |  | 0.004 |
| No | 11,693 | 11297 (96.6) | 396 (3.4) | 1 |  |
| Yes | 477 | 449 (94.1) | 28 (5.9) | 1.78 (1.17-2.59) |  |
| **Low 5 min Apgar score (<7)<sup>b</sup>** |  |  |  |  | <0.001 |
| No | 11,951 | 11555 (96.7) | 396 (3.3) | 1 |  |
| Yes | 219 | 191 (87.2) | 28 (12.8) | 4.28 (2.79-6.33) |  |
| **Stillbirth** |  |  |  |  | <0.001 |
| No | 12,101 | 11686 (96.6) | 415 (3.4) | 1 |  |
| Yes | 91 | 78 (85.7) | 13 (14.3) | 4.69 (2.47-8.22) |  |
| **Neonatal factors** |  |  |  |  |  |
| **Birth weight (Kg)<sup>c</sup>** |  |  |  |  | <0.001 |
| Mean (SD) |  | 3.0 (0.4) | 2.3 (0.5) | - |  |
| Normal (2.5-4) | 10,870 | 10696 (98.4) | 174 (1.6) | 1 |  |
| Low (<2.5) | 1156 | 903 (78.1) | 253 (21.9) | 17.22 (14.05-21.16) |  |
| Macrosomia (>4) | 156 | 156 (1.3) | 0 (0.0) | 0 (0.0) |  |
| **Easily recognisable congenital malformation** |  |  |  |  | 0.225 |
| No | 12,060 | 11634 (96.5) | 426 (3.5) | 1 |  |
| Yes | 132 | 130 (98.5) | 2 (1.5) | 0.42 (0.07-1.32) |  |
| **Postnatal outcomes** |  |  |  |  |  |
| **Neonatal admission** |  |  |  |  | 0.011 |
| No | 11,713 | 11312 (96.6) | 401 (3.4) | 1 |  |
| Yes | 479 | 452 (94.4) | 27 (5.6) | 1.69 (1.10-2.47) |  |
| **Mortality within 28 days<sup>d</sup>** |  |  |  |  | <0.001 |
| Alive | 11,959 | 11562 (96.7) | 397 (3.3) | 1 |  |
| Death | 142 | 124 (87.3) | 18 (12.7)<sup>e</sup> | 4.23 (2.55-5.70) |  |

CI = Confidence intervals; KG = Kilograms; SD = Standard deviation
a) N=2 missing data; b) N=22 missing data; c) N=10 missing data; d) For live-born neonates only

**Figure 2.**
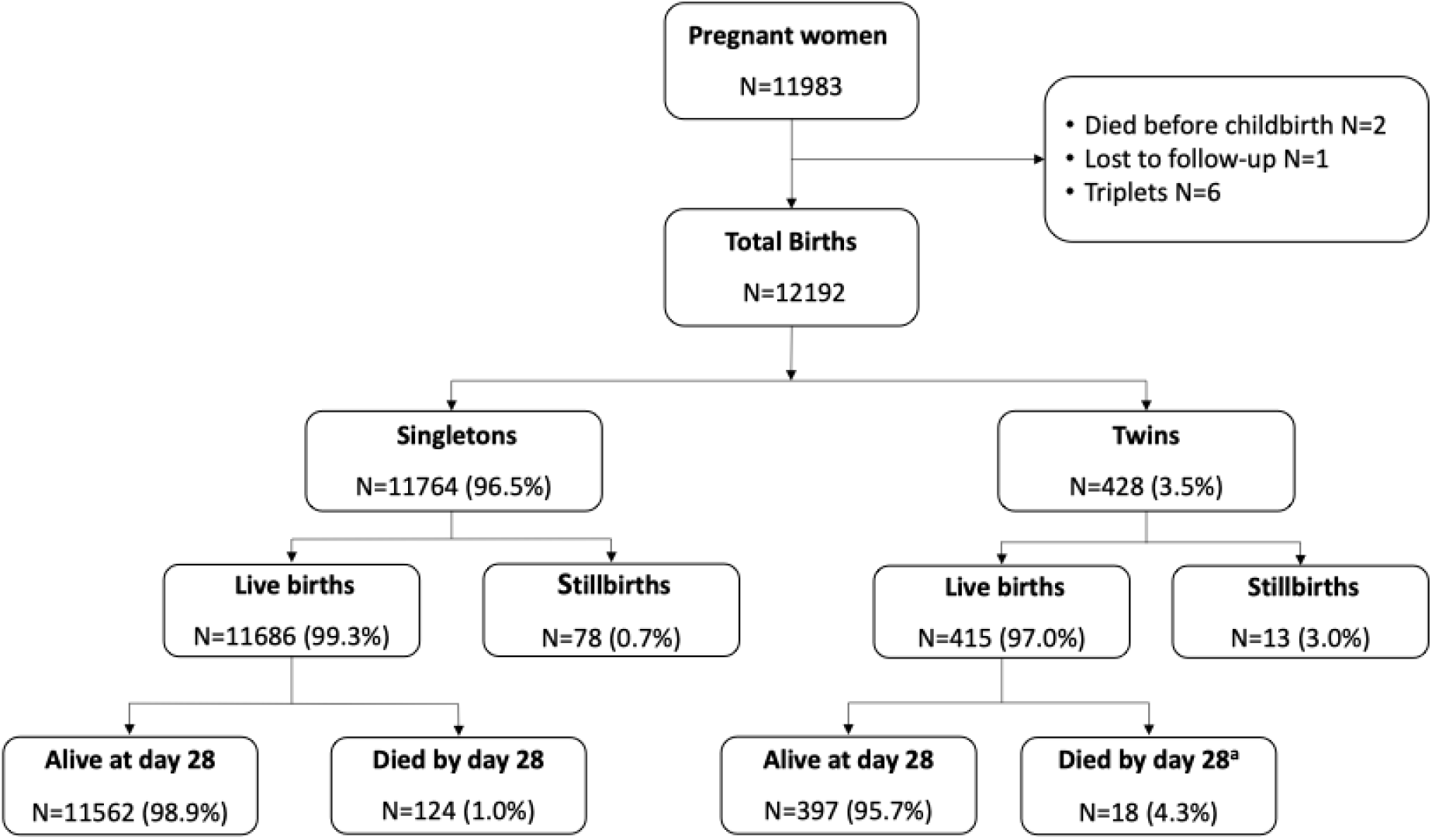
Participants and perinatal outcomes for singleton and twins born at health facilities in The Gambia and Burkina Faso. a) Of live-born twins who died, 66.7% (12/18) had the same mother (twin pairs) and 33.3% (6/18) had different mothers.

## DISCUSSION

This large health-facility based study underlines the importance of optimising management of twin pregnancies and deliveries in West Africa, with twins making a substantial contribution to poor perinatal and neonatal outcomes such as intrapartum stillbirths, neonatal mortality, intra-partum related asphyxia, LBW and hospital admission. We identified significant sub-regional variation in twinning rates in West Africa, being higher in Burkina Faso compared to The Gambia. Maternal factors associated with twinning in our cohort included age, parity and ethnicity.

The overall twinning rate in our West African cohort (35 per 1000 total births) is similar to findings from other health facility-based studies in the region, such as Nigeria (32.5 per 1000 total births)(23) and Ghana (33.4 per 1000 total births),(24), or countries from other Sub-Saharan regions such as Ethiopia (37.7 per 1000 total births)(25). These rates are considerably higher than population-based surveillance studies of twinning across SSA (17 per 1000 total births),(2) as well as in rural Gambia (15– 16.7 per 1000 births),(7, 26) and Burkina Faso (19.7-29.6 per 1000 livebirths).(2, 27) This disparity may be attributed to under-reporting of twin births in population-based studies, which typically rely on maternal recall of complete birth histories and often exclude stillbirths.(27) In contrast, our study prospectively collected data on all pregnancy outcomes within a clinical trial setting, providing a more accurate estimate of twinning prevalence among women delivering at West African health facilities. We also identified intra-regional variation in twinning prevalence, with a higher rate in Burkina Faso compared to The Gambia. Such regional variation is well documented, with West and Central African countries showing higher twinning rates compared to East and Southern Africa (2) and with substantial variability both between and within different West African countries.(2, 23) Our findings may also be partially explained by a higher proportion of women with parity ≥4 in the Burkina Faso sub-cohort and genetic differences present in varying ethnicities, which is speculated to be important in driving twinning in African populations.(2)

With a four-fold increased odds of neonatal mortality, the contribution of twins to neonatal mortality in our cohort was significant (12.5%) and is consistent with previous studies. (7, 27) The contribution of twins to neonatal mortality is now well evidenced from all African regions (2, 5, 27, 28, 29) and underlines the urgent need for global policy and programmes to prioritise early twin detection, enhanced antenatal monitoring and intrapartum management as a strategy to reduce avoidable stillbirths, IRA, and neonatal deaths. We had similar observations for intrapartum stillbirths. The prevalence of intrapartum stillbirth in our health facility-born twin cohort (13%) aligns with rates reported in Nigerian health facilities (10.8%)(23) and is substantially lower than rates reported from East African twin studies (30%).(28) The association between twin delivery and low Apgar scores observed in our study and others(30) suggests that IRA is an important factor in the development of intrapartum stillbirth and neonatal mortality for twins. Foetal malpresentation (e.g., breech or transverse lie) of one or both twins occurs in up to 50% of twin deliveries in similar West African settings (23) and may be an important event in the development of IRA, intrapartum stillbirth and neonatal mortality in our twin cohort, although specific data on this is not available in our study.

Other adverse outcomes which were more prevalent among twins in our cohort included LBW, birth by caesarean section and hospital admission. Our estimate of a 17-fold increased odds of LBW in twins is consistent with findings from other settings, (3, 4, 28, 31) with nearly half of all twin pregnancies resulting in LBW in East Africa.(28) LBW typically occurs in twins due to premature delivery, growth restriction, or both. However, we are unable to comment on the underlying cause of LBW in our cohort due to the lack of reliable gestational age data, an ongoing challenge in settings without routine first trimester ultrasound scans. We also observed a 2-fold increased odds of twin neonates born by emergency caesarean section, reflecting likely intrapartum complications, also observed in other health-facility West African twin studies.(3, 4, 32)

Higher parity (≥3), maternal age 20-35yrs and specific maternal ethnicities were associated with increased odds of twinning in our study. Women aged <20yr had a lower odds of having twins compared to women aged 20-35yrs, and there was a trend towards lower twinning rates for women aged >35y. This pattern is consistent with other SSA studies (2, 5) and could be attributed to age-related changes in maternal gonadotrophin levels and decline in ovarian function at the end of the reproductive cycle.(33) Our finding that twins were more likely to be born to women with increasing parity is consistent with previous studies from SSA,(2) including The Gambia,(7) and the Indian sub-continent.(34) Similar to these studies, we identified that multiparous women have the highest odds of twins, independent of maternal age, and consistent with previous evidence that maternal age and parity have independent positive associations with twin births.(5) Gambian ethnicities did not show any increased risk and had overall lower odds of twinning compared to the Mossi ethnic group in Burkina Faso. There is conflicting data on the influence of ethnicity on twinning in Gambian women, with reduced twinning rates in rural Mandinka populations historically reported,(26) yet a more recent population-based rural study finding no ethnic predisposition.(7) Our findings suggest that early (first trimester) targeted screening of multiparous women may enable early detection of twin pregnancies and provide a feasible strategy leading to optimised antenatal care, including foetal growth monitoring, promotion of maternal nutrition, and detection of obstetric complications known to be more prevalent in twin pregnancies (e.g., hypertensive disorders of pregnancy).(28)

## Conclusion

Twins represent a highly vulnerable neonatal population in The Gambia and Burkina Faso, with among the highest prevalence rates globally and a significantly increased risk of LBW, intrapartum stillbirth, IRA and neonatal mortality. Multiparous women from specific ethnic groups are at higher risk of twinning. There is an urgent need to implement systematic screening strategies for early detection of twin pregnancies, paired with high quality and context specific antenatal and intrapartum management. Strengthening these services is essential if the SDG target of reducing preventable neonatal mortality is to be achieved in West African countries by 2030.

## Data Availability

Data may be obtained from a third party and are not publicly available. The clinical data has been collected following provision of informed consent under the prerequisite of strict participant confidentiality. Qualified researchers may request access with the Gambia Government/MRC Joint Ethics Committee. The review process and release of data will be facilitated by MRC Unit The Gambia (http://www.mrc.gm/) through the Head of Governance at MRCG. Access will not be unduly restricted.

## DECLARATIONS

### Ethical approval

The parent trial was approved by The Gambia Government/Medical Research Council Unit The Gambia (MRCG) Joint Ethics Committee, the Comité d’Ethique pour la Recherche en Santé (CERS) and the Ministry of Health of Burkina Faso, and the LSHTM Ethics Committee. All women provided written informed consent during antenatal care visits and were free to withdraw at any time.

### Authors’ contributions

AR conceptualised this study with input from UdA, HB, AT, BC, JDB. Data collection was co-ordinated by BC, JDB and PregnAnZI-2 field teams in The Gambia and Burkina Faso. TR performed the analysis with full access to the data. JCJ drafted the initial manuscript with input from HB and AR. All authors contributed to the final version. AR gave oversight to the work as guarantor and accepts full responsibility for finished work and controlled the decision to publish.

## Acknowledgements

We acknowledge the project managers and all the field, data-management, and laboratory teams at MRCG and CRUN for providing support to conduct of the main trial. We also thank the leadership boards and staff at the study sites for their support. We are grateful to all the mothers and their neonates who participated to this study.

## REFERENCES

1. Smits J, Monden C. Twinning across the Developing World. PLOS ONE. 2011;6(9):e25239.

2. Gebremedhin S. Multiple Births in Sub-Saharan Africa: Epidemiology, Postnatal Survival, and Growth Pattern. Twin Research and Human Genetics. 2015;18(1):100–7.

3. Obiechina N, Okolie V, Eleje G, Okechukwu Z, Anemeje O. Twin versus singleton pregnancies: the incidence, pregnancy complications, and obstetric outcomes in a Nigerian tertiary hospital. Int J Womens Health. 2011;3:227–30.

4. Santana DS, Silveira C, Costa ML, Souza RT, Surita FG, Souza JP, et al. Perinatal outcomes in twin pregnancies complicated by maternal morbidity: evidence from the WHO Multicountry Survey on Maternal and Newborn Health. BMC Pregnancy and Childbirth. 2018;18(1):449.

5. Monden CWS, Smits J. Mortality among twins and singletons in sub-Saharan Africa between 1995 and 2014: a pooled analysis of data from 90 Demographic and Health Surveys in 30 countries. The Lancet Global Health. 2017;5(7):e673–e9.

6. Goetghebuer T, Ota MOC, Kebbeh B, John M, Jackson-Sillah D, Vekemans J, et al. Delay in Motor Development of Twins in Africa: A Prospective Cohort Study. Twin Research and Human Genetics. 2003;6(4):279–84.

7. Miyahara R, Jasseh M, Mackenzie GA, Bottomley C, Hossain MJ, Greenwood BM, et al. The large contribution of twins to neonatal and post-neonatal mortality in The Gambia, a 5-year prospective study. BMC Pediatrics. 2016;16(1):39.

8. Bjerregaard-Andersen M, Lund N, Jepsen FS, Camala L, Gomes MA, Christensen K, et al. A prospective study of twinning and perinatal mortality in urban Guinea-Bissau. BMC Pregnancy and Childbirth. 2012;12(1):140.

9. Morales-Suárez-Varela MM, Bech BH, Christensen K, Olsen J. Coffee and smoking as risk factors of twin pregnancies: the Danish National Birth Cohort. Twin Res Hum Genet. 2007;10(4):597–603.

10. Olusanya BO. Perinatal Outcomes of Multiple Births in Southwest Nigeria. Journal of Health, Population and Nutrition. 2011;29(6):639–47.

11. Onwuzuruike B, Onah H. Caesarean section in twin pregnancies in Enugu, Nigeria. International Journal of Medicine and Health Development. 2004;9(1):8–11.

12. Liu L, Chu Y, Oza S, Hogan D, Perin J, Bassani DG, et al. National, regional, and state-level allcause and cause-specific under-5 mortality in India in 2000–15: a systematic analysis with implications for the Sustainable Development Goals. The Lancet Global Health. 2019;7(6):e721–e34.

13. Hanson C, Munjanja S, Binagwaho A, Vwalika B, Pembe AB, Jacinto E, et al. National policies and care provision in pregnancy and childbirth for twins in Eastern and Southern Africa: A mixed-methods multi-country study. PLoS Med. 2019;16(2):e1002749.

14. Camara B, Bognini JD, Nakakana UN, Some AM, Jagne I, Tahita MC, et al. Pre-delivery administration of azithromycin to prevent neonatal sepsis and death: a phase iii double-blind randomized clinical trial (PregnAnZI-2 trial). 2022. 2022;9(1):11.

15. Roca A, Camara B, Bognini JD, Nakakana UN, Somé AM, Beloum N, et al. Effect of Intrapartum Azithromycin vs Placebo on Neonatal Sepsis and Death: A Randomized Clinical Trial. Jama. 2023;329(9):716–24.

16. Gambia Bureau of Statistics - GBoS, ICF. The Gambia Demographic and Health Survey 2019-Banjul, The Gambia: GBoS/ICF; 2021.

17. Institut National de la Statistique et de la Démographie, The DHS Program. Burkina Faso Enquête Démographique et de Santé 2021. Burkina Faso et Rockville, Maryland, USA: INSD et ICF; 2023.

18. United Nations Inter-agency Group for Child Mortality Estimation. Levels & Trends in Child Mortality: Report 2022. New York: UNICEF; 2023 2023.

19. United Nations Inter-agency Group for Child Mortality Estimation. Never Forgotten: The situation of stillbirth around the globe. New York: UNICEF; 2023.

20. Camara B, Bognini JD, Nakakana UN, Some AM, Jagne I, Tahita MC, et al. Pre-delivery administration of azithromycin to prevent neonatal sepsis and death: a phase iii double-blind randomized clinical trial (PregnAnZI-2 trial). International Journal of Clinical Trials. 2022;9(1):34-.

21. (WHO) WHO. ICD-11 International Classification of Diseases 11th Revision The global standard for diagnostic health information WHO; 2019.

22. Westreich D, Greenland S. The table 2 fallacy: presenting and interpreting confounder and modifier coefficients. Am J Epidemiol. 2013;177(4):292–8.

23. Akaba GO, Agida TE, Onafowokan O, Offiong RA, Adewole ND. Review of twin pregnancies in a tertiary hospital in Abuja, Nigeria. J Health Popul Nutr. 2013;31(2):272–7.

24. Mosuro AA, Agyapong AN, Opoku-Fofie M, Deen S. Twinning Rates in Ghana. Twin Research. 2001;4(4):238–41.

25. Tilahun T, Araya F, Tura G. Incidence and risk factors of twin pregnancy at Jimma University Specialized Hospital, Southwest Ethiopia. Epidemiology (sunnyvale). 2015;5(188):2161–1165.1000188.

26. Jaffar S, Jepson A, Leach A, Greenwood A, Whittle H, Greenwood B. Causes of mortality in twins in a rural region of The Gambia, West Africa. Ann Trop Paediatr. 1998;18(3):231–8.

27. Jahn A, Kynast-Wolf G, Kouyaté B, Becher H. Multiple pregnancy in rural Burkina Faso: frequency, survival, and use of health services. Acta Obstet Gynecol Scand. 2006;85(1):26–32.

28. Getachew T, Negash A, Debella A, Yadeta E, Lemi M, Balis B, et al. Prevalence and adverse outcomes of twin pregnancy in Eastern Africa: a systematic review and meta-analysis. BMC Pregnancy and Childbirth. 2024;24(1):169.

29. Isaacson A, Diseko M, Mayondi G, Mabuta J, Davey S, Mmalane M, et al. Prevalence and outcomes of twin pregnancies in Botswana: a national birth outcomes surveillance study. BMJ Open. 2021;11(10):e047553.

30. Santana DS, Cecatti JG, Surita FG, Silveira C, Costa ML, Souza JP, et al. Twin Pregnancy and Severe Maternal Outcomes: The World Health Organization Multicountry Survey on Maternal and Newborn Health. Obstetrics & Gynecology. 2016;127(4):631–41.

31. Assunção RA, Liao AW, Brizot Mde L, Krebs VL, Zugaib M. Perinatal outcome of twin pregnancies delivered in a teaching hospital. Rev Assoc Med Bras (1992). 2010;56(4):447–51.

32. Onyiriuka AN. Twin delivery: comparison of incidence and foetal outcome in two health institutions in Benin City, Nigeria. Nig Q J Hosp Med. 2009;19(1):1–5.

33. Bortolus R, Parazzini F, Chatenoud L, Benzi G, Bianchi MM, Marini A. The epidemiology of multiple births. Hum Reprod Update. 1999;5(2):179–87.

34. Satija M, Sharma S, Soni R, Sachar R, Singh G. Twinning and its correlates: community-based study in a rural area of India. Human Biology. 2008;80(6):611–21.

