## Supplementary Table 1 for "Clinical epidemiology of twin deliveries in The Gambia and Burkina Faso: Secondary analyses of PregnAnZI-2 clinical trial data"

Supplementary table 1. Participant characteristics stratified by country

|  | **Total** | **Burkina Faso** | **Gambia** | ***p-value*** |
| --- | --- | --- | --- | --- |
|  | n=12192 | n=5369 | n=6823 |  |
| Twin deliveries (n, %) | 428 (3.5) | 244 (57.0) | 184 (43.0) | <0.001 |
| Maternal age (years) |  |  |  |  |
| Mean (SD) | 26.8 (6.1) | 26.3 (6.3) | 27.2 (6.0) |  |
| 16-19 | 1526 (12.5) | 888 (16.5) | 638 (9.4) | <0.001 |
| 20-35 | 9540 (78.2) | 4032 (75.1) | 5508 (80.7) |  |
| >35 | 1126 ( 9.2) | 449 ( 8.4) | 677 (9.9) |  |
| Maternal ethnicity (n,%) |  |  |  | <0.001 |
| Mossi | 4942 (40.5) | 4942 (92.0) | 0 (0.0) |  |
| Gurunsi | 351 (2.9) | 351 (6.5) | 0 (0.0) |  |
| Mandinka | 2722 (22.3) | 0 (0.0) | 2722 (39.9) |  |
| Fula | 1296 (10.6) | 69 (1.3) | 1227 (18.0) |  |
| Wollof | 1046 ( 8.6) | 0 ( 0.0) | 1046 (15.3) |  |
| Jola | 887 (7.3) | 0 (0.0) | 887 (13.0) |  |
| Other | 948 ( 7.8) | 7 ( 0.1) | 941 (13.8) |  |
| Maternal parity (n,%) |  |  |  | <0.001 |
| Primipara | 2756 (22.6) | 1053 (19.6) | 1703 (25.0) |  |
| Parous 2 | 2245 (18.4) | 846 (15.8) | 1399 (20.5) |  |
| Parous 3 | 1970 (16.2) | 793 (14.8) | 1177 (17.3) |  |
| Parous >=4 | 5220 (42.8) | 2676 (49.9) | 2544 (37.3) |  |
| Conceived during rainy season (n,%) | 4616 (37.9) | 2062 (38.5) | 2554 (37.5) | 0.297 |

SD = Standard deviation
